# Moving Past Tonsil Position: Craniocervical Junction Crowding Shapes Cerebrospinal Fluid Effective Motility in Chiari I Malformation

**DOI:** 10.64898/2026.07.28.26358336

**Authors:** Helia Hosseini, Amir H. Shaker, Connor A. Sierra, Wan-Yun Shen, Zhouqiao Zhao, Farrell Landwehr, Aristeidis Sotiras, Joshua S. Shimony, Bryn A. Martin, David D. Limbrick, Jennifer M. Strahle, Arash Nazeri

## Abstract

Chiari I malformation (CM-I) is conventionally defined by cerebellar tonsil position, yet tonsil position is an indirect surrogate for the anatomic obstruction that impairs cerebrospinal fluid (CSF) flow across the craniocervical junction (CCJ). We hypothesized that CCJ crowding, quantified as subarachnoid space narrowing at the foramen magnum and C1, would explain CSF flow impairment more directly than tonsil position. Using non-invasive low b-value diffusion-weighted MRI (low-b dMRI), we quantified effective CSF motility, indexed by mean pseudo-diffusivity (MΨ), across the upper cervical spine, CCJ, and posterior fossa. Voxel-wise CCJ CSF pseudo-diffusion spatial statistics were integrated with CSF Waterways atlas-based regional analyses. We applied this approach in 81 pediatric and adult participants with CM-I to determine how CCJ structural features shape regional CSF dynamics and clinical outcomes. Voxel-wise analyses revealed that crowding at the foramen magnum was the dominant structural determinant of reduced intracranial CSF effective motility across the CCJ, basilar cisterns, and fourth ventricular outflow pathways (family-wise error corrected *p* < 0.05). While lower tonsil position and C1 level crowding were also associated with reduced CSF effective motility across the CCJ and fourth ventricular outflow pathways, but their associations within the basilar cisterns were spatially restricted to regions adjacent to the Liliequist membrane. Atlas-based region-of-interest analyses confirmed that greater foramen magnum crowding was associated with lower MΨ across multiple basilar cisterns, but with higher MΨ in the ventral spinal CSF compartment. Mediation analyses indicated that CCJ crowding at the foramen magnum and C1 accounted for the majority of the relationship between tonsil position and reduced CSF motility in the basilar cisterns. Multivariate MΨ profiles across the CSF regions identified data-driven foramen magnum crowding thresholds of 69.5% and 77.5%, stratifying patients into mild, moderate, and severe physiological crowding groups. Exploratory analyses linked lower pre-operative CSF MΨ to greater pain-related functional impairment, reduced cognitive function, and a higher likelihood of subsequent decompression surgery. Together, these findings demonstrate that CCJ crowding, particularly at the foramen magnum, exerts a quantifiable, region-specific impact on CSF effective motility in CM-I, and that low-b dMRI provides a sensitive, complementary marker of CSF flow impairment. This integrative CCJ structural and CSF flow imaging framework establishes a mechanistic link between CCJ anatomy, CSF dynamics, and symptom burden, offering a scalable tool for phenotyping CM-I and informing clinical decision-making.

## Introduction

Chiari I malformation (CM-I) encompasses a spectrum of congenital or acquired hindbrain and craniocervical junction (CCJ) structural abnormalities defined by caudal displacement of the cerebellar tonsils below the foramen magnum^1^. A key structural consequence of cerebellar tonsillar descent is crowding at the foramen magnum and posterior fossa, often manifested by effacement of the surrounding cerebrospinal fluid (CSF) spaces^2,3^. The resulting crowding and structural narrowing at the foramen magnum can alter CSF flow dynamics across the foramen magnum, basilar cisterns, and ventricular system, disrupting intracranial pressure regulation and secondarily driving aberrant motion of the tonsils and CCJ^4^. Classic clinical features of CM-I, such as cough- or Valsalva-induced suboccipital headache, are thought to arise from sudden increases in intracranial pressure due to impaired CSF outflow at the foramen magnum, resulting from descent of the cerebellar tonsils and consequent intracranial dural distention^5,6^.

The pathophysiology of CM-I reflects the interaction between the CCJ structural constraint and CSF flow dynamics and cannot be adequately captured by a single parameter such as tonsil position^1,7^. Growing evidence suggests that CM-I associated symptoms, particularly headache, arise from impaired CSF flow limitation imposed at the CCJ, rather than low tonsil position alone^8–10^. Some have conceptualized CSF flow impairment in CM-I as a dorsal-to-ventral pattern of disease severity, with abnormalities described posterior to the cerebellar tonsils and dorsal spinal CSF column, at the fourth ventricular outlets, and, in more severe cases, within the ventral CCJ and upper cervical CSF spaces^6,11^. However, this proposed severity framework has not been quantitatively established and is based on qualitative imaging observations. Moreover, prior studies have often used tonsil position as the principal structural marker of CM-I severity, despite its poor correlation with symptom severity and surgical outcomes^12–14^. This reliance on tonsil position may obscure the structural features most directly linked to CSF flow impairment, underscoring the need for integrated analyses of CCJ anatomy and CSF dynamics to redefine the anatomic determinants of flow obstruction in CM-I.

Two-dimensional (2D) cardiac-gated phase-contrast MRI (PC-MRI) remains the conventional clinical approach for assessing CSF flow disturbances in CM-I^1^. Cardiac-gated PC-MRI studies have shown that patients with CM-I exhibit altered CSF velocities, flow jets, and asynchronous or bidirectional CSF flow near the CCJ^15–17^. Although 2D PC-MRI captures directional, time-resolved flow velocities across the cardiac cycle, it necessitates the selection of predetermined imaging planes, which may not accurately align with regions of disturbed flow in CM-I. Four-dimensional (4D) flow MRI offers volumetric assessment of multidirectional CSF velocities with expanded spatial coverage, yet its clinical utility remains limited by prolonged acquisition times and technical complexity. Notably, both 2D and 4D PC-MRI require prior specification of a velocity encoding (VENC) parameter that limits dynamic range—where high VENC reduces sensitivity to slow flow and low VENC increases aliasing in fast flow—and are inherently constrained in capturing complex vortical, bidirectional, and mixing flow patterns^18^. Finally, most studies employing 4D flow MRI rely on predefined regions of interest, even though the precise locations of flow disturbance in CM-I are often unknown. This reliance on subjective regions of interest placement may obscure critical pathophysiological features and highlights the need for more comprehensive, data-driven approaches. These methodological limitations may partly explain why 2D PC-MRI-derived CSF flow metrics have not demonstrated robust utility for guiding surgical management in CM-I^19^.

In contrast to PC-MRI, low-b diffusion-weighted MRI (low-b dMRI) can capture slow and complex CSF motion^18,20–23^. This is achieved using a pulsed gradient spin-echo sequence with rapid echo-planar imaging (EPI) readout, which takes advantage of the long T2 relaxation time of CSF^21^. Rather than relying on phase shifts, low-b dMRI measures changes in signal magnitude loss caused by pseudorandom fluid displacement within each voxel. The resulting mean pseudo-diffusivity (MΨ)—analogous to mean diffusivity at higher b-values—serves as a marker of effective CSF motility, capturing a spectrum of flow regimes including laminar, vortical, and mixing patterns^18,20,21,24^, including those mediated by cardiac pulsatility and other physiologic drivers. Rapid EPI acquisitions enable three-dimensional fast CSF flow mapping within the field of view. These properties make low-b dMRI complementary to PC-MRI for CSF flow mapping across the upper cervical spine, CCJ, and posterior fossa in CM-I, where altered and spatially heterogeneous CSF flow patterns are common and may not be fully characterized by bulk velocity changes across the cardiac cycle.

To directly interrogate how CCJ anatomy shapes CSF flow across the upper cervical spine, CCJ, and posterior fossa, we developed a tailored imaging and analytical framework based on low-b dMRI. We implemented a high-resolution, reduced field-of-view (ZOOMit) acquisition centered at the CCJ to enhance image fidelity and introduced a modified voxel-wise CSF flow analysis framework, CCJ-CSF pseudodiffusion spatial statistics (CCJ-CΨSS). In parallel, we used the CSF Waterways (CWW) atlas^21^ for region-of-interest analyses of CSF flow changes across spinal, ventricular, and basilar cistern compartments. Notably, CCJ crowding, quantified as the percentage of subarachnoid-space obliteration on cross-sectional reformats at the foramen magnum and C1-levels, was more strongly associated with CSF effective motility than tonsil position. Using this integrative structural-functional CSF flow imaging approach, we identified data-driven foramen magnum crowding thresholds that defined stepwise differences in regional CSF effective motility among patients with CM-I. Finally, low-b dMRI-derived MΨ captured information complementary to C2–C3 PC-MRI and was more strongly and independently associated with CCJ crowding than conventional PC-MRI flow measures. The overall study design and analytical workflow are summarized in **Fig. 1**.

**Fig. 1.**
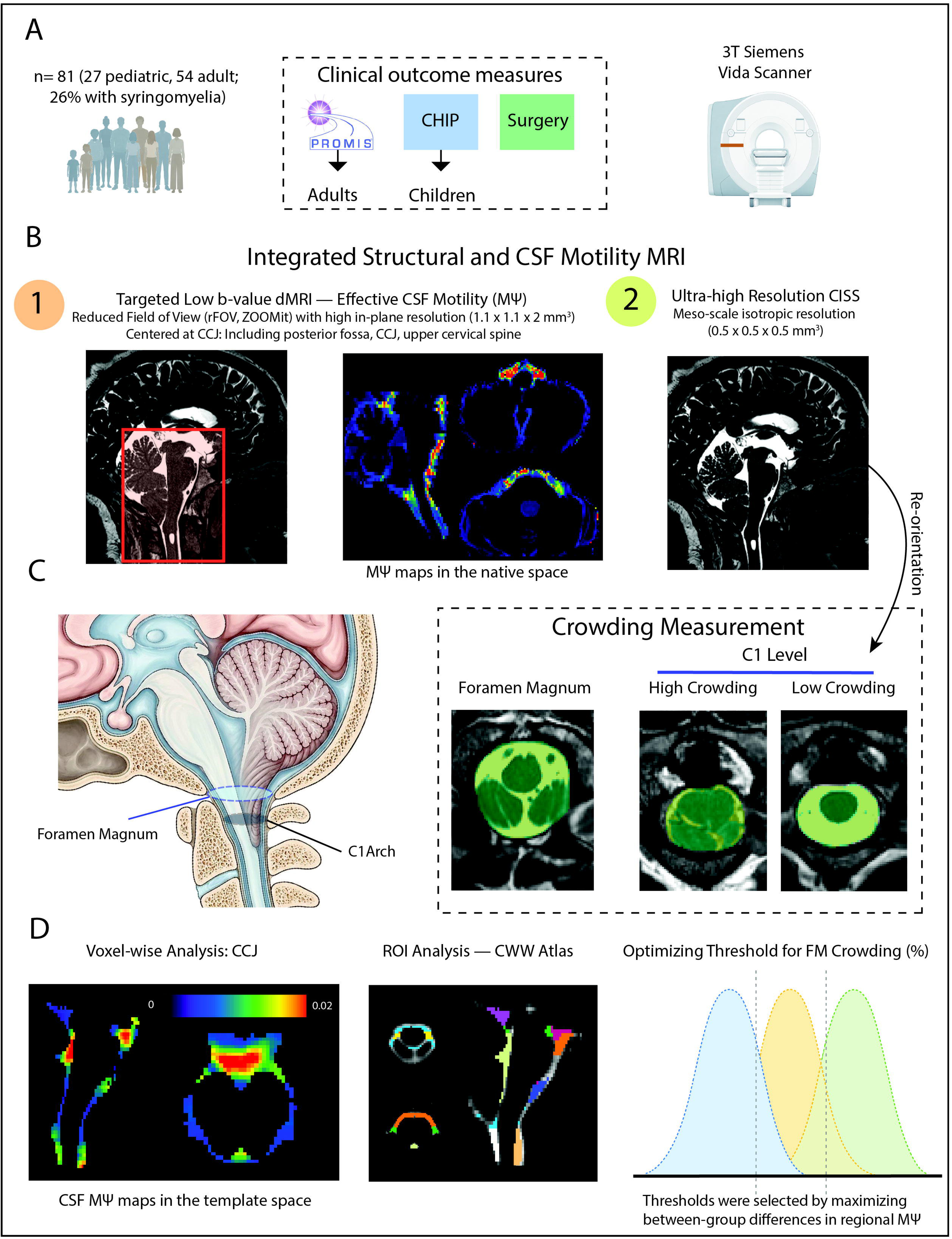
Overview of the study design, imaging acquisition, and analytical framework. **(A)** Study cohort, clinical assessments, and MRI acquisition. **(B)** Integrated structural and functional imaging using a (1) high resolution, reduced field-of-view (ZOOMit) low b-value diffusion MRI acquisition centered at the craniocervical junction to quantify effective CSF motility (M) and (2) ultra-high-resolution 3D CISS imaging for structural assessment. **(C)** Craniocervical junction crowding was quantified as the percentage of subarachnoid space obliteration at the foramen magnum and C1 on reoriented 3D CISS images. **(D)** Image analysis workflow. Low-b-value dMRI preprocessing and cranio-cervical junction CSF pseudo-diffusion spatial statistics (CCJ-CΨSS) were used to generate MΨ maps and perform voxel-wise analysis. CSF Waterways (CWW) atlas-based region-of-interest analyses were then performed, followed by data-driven identification of foramen magnum crowding thresholds. *Abbreviations: CSF, cerebrospinal fluid; MΨ, mean pseudo-diffusivity; CISS, constructive interference in steady state; CCJ, craniocervical junction;*.

## Methods

### Participants

This study is part of a larger single-site prospective cohort, Redefining Chiari Type I Malformation and its Impact on Brain Development (NIH P01NS131131; sub-project 5757), conducted at Washington University School of Medicine in collaboration with St. Louis Children’s Hospital and Barnes-Jewish Hospital. Individuals were recruited from affiliated clinical services. This study was reviewed and approved by the Institutional Review Board at the Washington University in St. Louis (IRB# 202310017). Written informed consent was obtained from all participants or their legal guardians. The study included pediatric and adult patients who had tonsillar ectopia or CM-I, defined by cerebellar tonsillar descent of at least 3 mm below the foramen magnum. Exclusion criteria included a diagnosis of CM type II–IV, tonsil descent secondary to other structural pathologies, prior posterior fossa decompression, contraindications to MRI, or unwillingness to participate. All participants completed patient-reported outcome measures as part of the broader cohort protocol.

### Clinical assessment

In adults, health-related quality of life (HRQoL) was quantified using the Patient-Reported Outcomes Measurement Information System (PROMIS), including pain interference (PI), physical function (PF), and cognitive function (CF). Scores are reported as standardized T-scores (mean□=□50, SD□=□10), with higher values reflecting greater pain interference or improved function depending on the domain^25^. In pediatric patients, HRQoL was assessed using the Chiari Health Index for Pediatrics (CHIP), capturing both physical (pain severity, pain frequent, and non-pain symptoms) and psychological domains, with higher scores indicating better quality of life^26^.

### MRI imaging

Participants were scanned on a 3 Tesla Siemens Magnetom Vida MRI scanner with a 64-channel head/neck coil at the Center for Clinical Imaging Research (CCIR), Mallinckrodt Institute of Radiology. For CSF flow mapping, we used a low-b dMRI sequence optimized to capture CSF motion across the basilar cisterns, craniocervical junction, and upper cervical spine. Images were acquired using an axial reduced field of view (ZOOMit) 2D echo-planar imaging (EPI) sequence using the following imaging parameters: TR/TE, 7100/94 ms; field of view (FOV), 180 × 72 mm^2^; voxel size, 1.1 × 1.1 × 2 mm^3^; slice number, 48. By selectively exciting and encoding only the targeted region, ZOOMit enables high spatial resolution with reduced susceptibility artifacts^27^. Each diffusion gradient had a unique combination of b-value and direction, ranging from 0 to 1500 s/mm^2^, yielding a total of 55 volumes. For structural imaging of the CCJ, a high-resolution mid-sagittal 3D constructive interference in steady state (CISS) sequence was used with the following parameters: TR, 4.78 ms; TE, 2.13 ms; flip angle, 30^°^; FOV, 200 × 200 mm^2^; slice number, 96; voxel size, 0.5 × 0.5 × 0.5 mm^3^. This balanced steady state free precession (b-SSFP) sequence is inherently flow-compensated and provides high signal-to-noise ratio in CSF, enabling detailed visualization of fine anatomical details within CSF spaces due to the high T2/T1 ratio characteristic of CSF^28,29^. Axial cardiac-gated PC-MRI with retrospective peripheral pulse gating was performed at the C2–C3 level, with a velocity encoding (VENC) of 16 cm/s. Acquisition parameters were as follows: TR, 70.1 ms; TE, 7.18 ms, flip angle, 10°; slice thickness, 4 mm; in-plane resolution, 0.6 × 0.6 mm^2^; FOV: 230 × 230 mm^2^; calculated phases, 40, acquisition time: 1:50.

### Structural analyses

A total of 81 participants had available cerebellar tonsil position measurements and CCJ low-b dMRI. Tonsil position was quantified on mid-sagittal structural images by measuring the perpendicular distance from the tip of cerebellar tonsils to McRae’s line, which was defined by a straight line drawn between basion and opisthion. Crowding at the CCJ in CM was assessed using high-resolution 3D-CISS images as follows. All CISS images underwent quality control, and those scans with substantial motion or other artifacts were excluded. Each CISS image was first reoriented to align the McRae’s line (basion–opisthion) horizontally, standardizing the CCJ plane. The foramen magnum was then manually segmented. Voxels within this mask were input to a two-tissue segmentation algorithm (Atropos^30^, part of ANTs^31,32^) generating partial-volume maps for tissue and CSF. Crowding at the foramen magnum was quantified as the percentage of non-CSF voxels within the segmented region, consistent with prior studies^12,33^. A similar approach was used to assess crowding at the level of the C1 following reorientation of the CISS images along the anterior and posterior arches of the C1. After quality control, crowding measurements at both levels were available for 77 participants based on image availability.

### Low-b dMRI preprocessing and CSF pseudo-diffusion spatial statistics (CΨSS)

At low b-values, the measured pseudo-diffusion tensor **Ψ** can be interpreted as comprising two principal components: **D**, which reflects intrinsic molecular diffusivity together with random, disordered CSF motion, and **V**, the covariance matrix of the three-dimensional velocity probability distribution, which captures ordered linear CSF motion. Importantly, within CSF, the contribution from random, disordered motion may be substantially larger than intrinsic molecular diffusivity alone. Accordingly, Ψ can be expressed as the sum of D and V, with τ_d_ denoting the diffusion time^18,20^:

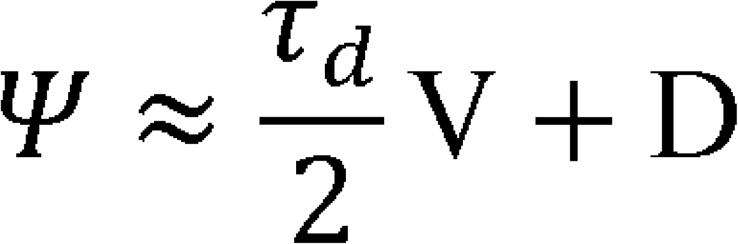

Assuming that the physical properties of CSF (e.g., temperature and viscosity) remain unchanged, changes in MΨ derived from the Ψ tensor can be interpreted as reflecting changes in effective CSF motility. Specifically, MΨ captures intravoxel dispersion of CSF velocities arising from laminar (ordered) and random (disordered) flow^18,20^. Overall, this framework is conceptually analogous to Aris-Taylor dispersion, in which apparent diffusivity is enhanced by the variance of velocity distributions across streamlines produced by local velocity gradients^34,35^.

To derive MΨ maps from the diffusion-weighted data and perform group-level analyses of CSF flow changes at the CCJ, we applied a preprocessing and CΨSS postprocessing workflow^21^. To correct for susceptibility-induced distortions and eddy currents, a single b0 volume was extracted from each phase-encoding direction (A>P and P>A). Off-resonance field estimation was then performed with FSL’s *topup*^36^, using the corresponding acquisition parameters. The resulting corrected b0 image was then averaged across volumes and skull-stripped using the brain extraction tool (BET) to generate a brain mask^37^. Where necessary, the mask was manually edited using ITK-SNAP (v.4.0.2)^38^ to include the spinal canal. Finally, eddy current-induced distortions and subject motion were corrected using FSL’s *eddy*^39^.

The multi-b-value diffusion MRI dataset (b = 0-1500 s/mm^2^) was then divided into low and high b-value subsets. The low b-value subset (b = 0-350 s/mm^2^) included 36 volumes (7 b0 images and 29 diffusion-weighted volumes) and was used to fit a diffusion tensor model, from which mean MΨ maps were derived as a surrogate of CSF effective motility. The high b-value subset (b ≥ 650 s/mm^2^) included 22 volumes (7 b0 images and 15 diffusion-weighted volumes) and was used to fit a separate diffusion tensor model to generate fractional anisotropy maps. The resulting low-b mean diffusivity (MΨ) and high-b fractional anisotropy maps were then fed into the Atropos^30^ to derive partial volume estimates of CSF and white matter using a two-tissue model (**Supplemental Fig. S1**). Similar to gray matter-based spatial statistics^40,41^, partial volume estimates were combined to synthesize pseudo-T1-weighted images, using gray matter, white matter, and CSF probability maps all derived from diffusion data. MΨ maps were thresholded to exclude physiologically implausible values. Study-specific CCJ templates were then constructed using both MΨ and pseudo-T1-weighted images as inputs to the *antsMultivariateTemplateConstruction2.sh* workflow. CSF probability maps were subsequently registered to the common template space. To minimize partial volume effects, the final CSF mask was created by retaining only those voxels where the CSF fraction exceeded 0.8 in at least 75% of the subjects. Voxels with CSF fraction less than 0.8 were then filled with the average of the surrounding satisfactory CSF voxels. Finally, spatial smoothing was then applied within the resulting CSF mask using the AFNI’s *3dBlurInMask* function^42^, with a full width at half maximum value of 3 mm.

### Region-of-interest analysis

To investigate regional CSF flow alterations, we performed of region-of-interest (ROI) analysis using the CSF Waterway (CWW) atlas—a data-driven parcellation that captures the intrinsic covariance structure of intracranial CSF flow^21^. The cisternal and ventricular CSF regions included in the CWW atlas were: CWW-1, foramen of Luschka; CWW-2, cisterna magna, cervicomedullary cistern, and cerebellopontine angles; CWW-3, superior premedullary cistern; CWW-4, chiasmatic cistern; CWW-5, prepontine cistern; CWW-6, perimesencephalic cisterns; CWW-9, supracerebellar and velum interpositum cisterns; and CWW-10, trans-ventricular pathway encompassing the third and fourth ventricles. The atlas was nonlinearly registered to a study-specific template using the ANTs^31,32^. Additionally, two ROIs corresponding to the dorsal and ventral subarachnoid spaces of the upper cervical spinal canal, extending from the CCJ to the C2–C3 level, were manually delineated in template space using ITK-SNAP. Mean MΨ values were then extracted from each CWW parcel using FSL’s *fslstats* tool.

### Outlier assessment

CSF MΨ maps in template space were converted to voxel-wise z-scores, and clusters with |z| > 3.5 and size > 50 voxels were identified. To assess potential underlying structural etiologies or incidental findings associated with aberrant CSF flow patterns, structural images of affected participants were reviewed. Based on these findings, the entire dataset was subsequently examined to identify similar structural abnormalities across participants. All image evaluations were performed by a board-certified neuroradiologist (A.N.).

### Quantification of spinal CSF dynamics at the C2–C3 level using phase-contrast MRI

Manual segmentation of the spinal canal and cord was performed on rephased magnitude images using ITK-SNAP. The spinal subarachnoid space was delineated as the region surrounding the spinal cord at the C2–C3 level. Prior to flow quantification, phase images were corrected for background phase offsets. For these spinal subarachnoid spaces the following metrics were extracted from PC MR images: (1) *peak systolic velocity*, defined as the maximum caudal CSF flow velocity during systole within the ROI; (2) *peak diastolic velocity*, defined as the maximum cranial CSF flow velocity during diastole within the ROI; (3) *stroke volume* was calculated as half the absolute area under the ROI-averaged CSF velocity–time curve across the cardiac cycle, multiplied by the cross-sectional area of the spinal subarachnoid space at C2–C3.

### Statistical analysis

Voxel-wise associations between CSF MΨ and anatomical markers of CM-I severity (tonsil position and CCJ crowding metrics) were assessed using FSL’s *randomise* with 5,000 permutations and threshold-free cluster enhancement for family-wise error (FWE) correction^43^. Separate general linear models were constructed for tonsil descent, CCJ crowding at both levels of foramen magnum and C1, adjusting for age and sex. Results were considered significant at FWE-corrected p < 0.05.

For region-of-interest analysis, general linear models were applied to mean MΨ values extracted from each of the 10 CWW regions to examine associations with CM-I structural markers. Separate and combined models were computed using tonsil descent, and CCJ crowding at both levels of foramen magnum and C1 measures, with all models adjusted for age and sex. False discovery rate (FDR) correction was applied across regions, with significance defined at FDR-corrected *p* < 0.05. To assess whether CCJ crowding mediated the relationship between tonsil descent and CSF effective motility, we performed parallel mediation analyses (*manymome* package in R^44^) to evaluate whether crowding at foramen magnum and C1 mediate the association between tonsil position and regional CSF MΨ. These analyses were restricted to CWW regions of interest that showed significant associations with tonsil position. All variables were standardized, and linear models were fit for each mediator and outcome, adjusting for age and sex. Indirect effects were estimated using nonparametric bootstrap resampling (5,000 iterations) with confidence intervals, and FDR correction was applied across effects. Crowding was stratified into mild, moderate, and severe levels using two data-driven cut-points. These cut-points were simultaneously optimized via a joint grid search over all valid cut-point pairs to maximize between-group separation in the multivariate profile of CWW regional MΨ values, as assessed by MANOVA with Pillai’s trace. The optimization was constrained to ensure adequate group size and separation, with at least 15 participants per group and at least a 5% crowding difference between thresholds. Candidate cut-points were evaluated across the 15th to 85th percentiles of the foramen magnum crowding distribution in 1-percentile increments. To assess threshold stability, we repeated the procedure across 1,000 bootstrap samples and report the median cut-points with 90% confidence intervals. Fisher exact tests were used to assess whether incidental structural findings were significantly associated with participants exhibiting extreme MΨ values. All statistical analyses were conducted using R (v4.5.3) and Python (v3.10.9).

## Results

### Demographic and clinical characteristics

A total of 81 participants with a clinical diagnosis of CM-I were included in this study, of whom 33.3% were pediatric (< 18 years). The mean age of the overall cohort was 27.1 ± 13.7 years, and 74.1% were female. Among these 81 individuals, 21 had syrinx and 23 underwent surgery. Mean tonsillar descent was 9.4 ± 4.9 mm in the overall cohort, with mean values of 8.8 ± 6.0 mm and 9.7 ± 4.4 mm in pediatric and adult participants, respectively. Summary descriptives of participants are detailed in **Table 1**. Among 77 participants with complete structural measurements, tonsillar descent correlated moderately with foramen magnum crowding (r = 0.46, p < 0.001) and strongly with C1 crowding (r = 0.71, p < 0.001). Foramen magnum and C1 crowding were only moderately correlated (r = 0.42, p < 0.001), suggesting that they capture distinct aspects of CCJ crowding.

**Table 1.**
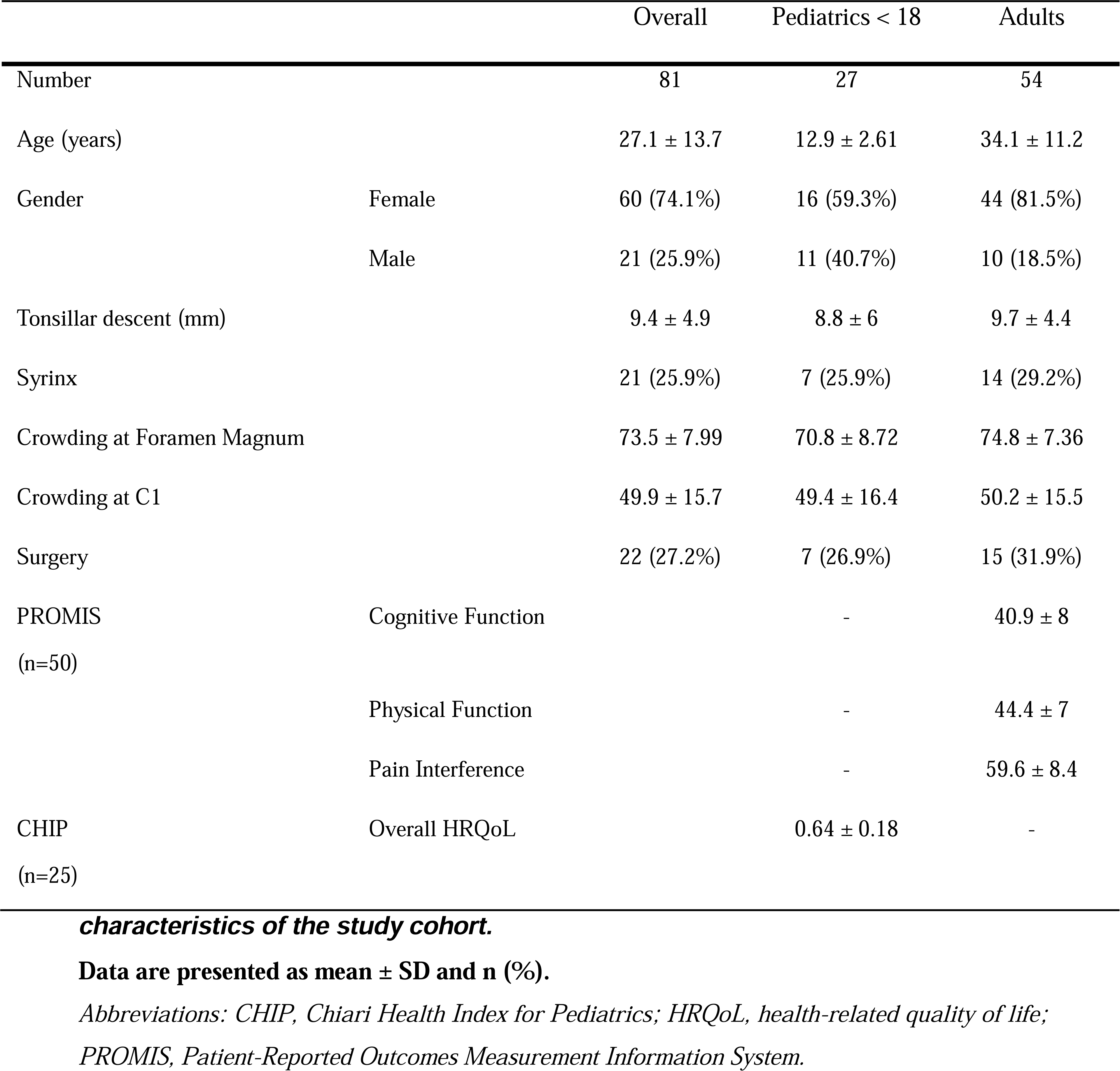
Demographic, clinical, and craniocervical junction structural.

### Voxel-wise Mapping Reveals Distinct Regional CSF Flow Alterations Associated with CCJ Crowding and Tonsil Position

Voxel-wise analysis revealed that greater CCJ crowding at the level of foramen magnum (n=77) was associated with lower CSF MΨ across the basilar cisterns, including the premedullary, prepontine, and ambient cisterns, interpeduncular cistern surrounding the Liliequist membrane (FWE-corrected *p* [*p_FWE_*] = 0.001), the fourth ventricular outflow pathways—the foramen of Magendie (*p_FWE_* = 0.02) and the foramina of Luschka (*p_FWE_* = 0.001), and the supracerebellar and velum interpositum cisterns (*p_FWE_* = 0.02; **Fig. 2A**). Compared to the more widespread associations between CSF MΨ and CCJ crowding at the level of foramen magnum in the basilar cisterns, CCJ crowding at the level of C1 (n=77) and tonsillar descent (n=81) were linked to more focal decreases in CSF MΨ, with significant clusters largely confined to the interpeduncular cistern surrounding the Liliequist membrane (*p_FWE_*= 0.01) and centered around the fourth ventricular outflow pathways (*p_FWE_*= 0.001; **Fig. 2B and 2C**). Overall, these spatial patterns suggest that CCJ crowding and tonsillar descent are associated with intracranial CSF flow impedance, reflected by diminished CSF motion in the basal cisterns.

**Fig. 2.**
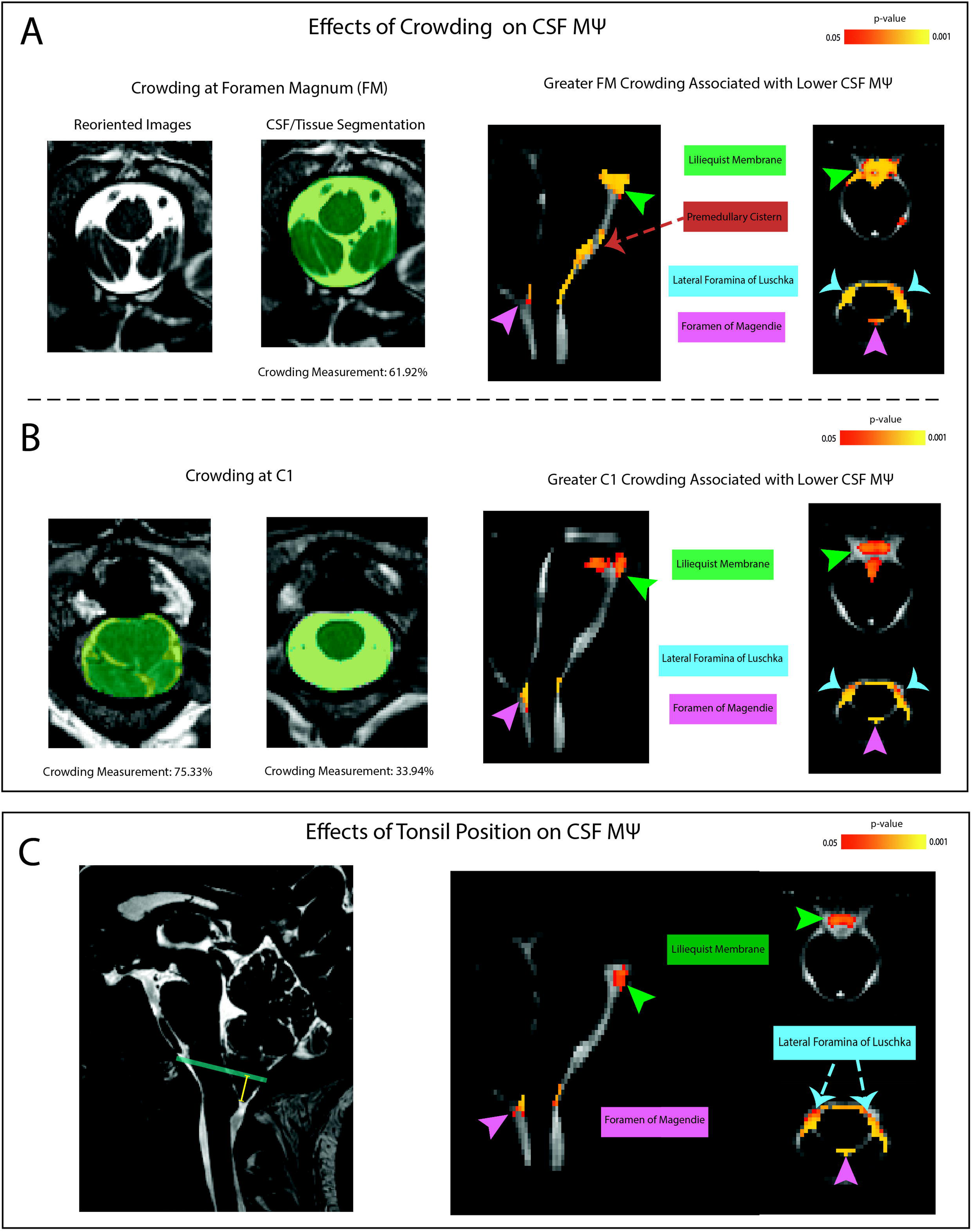
Voxel-wise mapping of CSF effective motility alterations associated with CCJ structural markers. **(A)** Greater tonsillar descent (n=81); measured as the perpendicular distance from the tip of cerebellar tonsils to McRae’s line, was associated with reduced CSF MΨ within interpeduncular cistern surrounding the Liliequist membrane (*p_FWE_* = 0.01) and the fourth ventricular outflow pathways (foramen of Magendie and foramina of Luschka; *p_FWE_*= 0.001). **(B)** Representative segmentation of the FM on reoriented high-resolution 3D-CISS images illustrates the quantification of crowding, defined as the percentage of non-CSF voxels. Voxel-wise analysis identified FM crowding (n=77) as the dominant structural determinant of reduced effective motility, showing widespread significant associations across the ventral basilar cisterns including the premedullary, prepontine and the interpeduncular cisterns (*p_FWE_* = 0.001), and the fourth ventricular outflow pathways (foramen of Magendie: *p_FWE_* = 0.02; foramina of Luschka: *p_FWE_* = 0.001). **(C)** Quantification of crowding at the C1-level demonstrated focal associations with reduced CSF MΨ. Significant clusters were primarily centered in the interpeduncular cistern (*p_FWE_* = 0.01) and the fourth ventricular outflow pathways (*p_FWE_* = 0.001). *Abbreviations: CSF, cerebrospinal fluid; CCJ, craniocervical junction; FM, foramen magnum; MΨ, mean pseudo-diffusivity; FWE, family-wise error*.

### Craniocervical Junction Crowding Mediates the Flow Attenuation Effects of Low Tonsil Position on the Basilar Cistern CSF Motility

Similar to voxel-wise analysis findings, region-of-interest analysis demonstrated a widespread negative association between CSF MΨ and crowding at the foramen magnum across multiple CWW regions^21^ within the posterior fossa and CCJ, accounting for age and sex. Specifically, outflow area of foramina of Luschka (CWW-1), cisterna magna (CWW-2), superior premedullary cistern (CWW-3), chiasmatic cistern (CWW-4), prepontine cistern (CWW-5), and perimesencephalic cisterns (CWW-6) all showed significant negative associations with greater crowding. By contrast, greater foramen magnum crowding was significantly associated with increased CSF MΨ in the ventral upper cervical spinal canal (*p_FDR_* < 0.05, **Fig. 3B and 3C**). Inverse associations of regional CSF MΨ with tonsil position and C1-level crowding were more anatomically restricted than those observed for foramen magnum crowding. For both measures, significant associations were confined to the foramina of Luschka outflow region (CWW-1), cisterna magna (CWW-2), and prepontine cistern (CWW-5); C1-level crowding showed an additional inverse association with MΨ in the third and fourth ventricles (CWW-10; pFDR < 0.05; **Fig. 3B**). Subgroup analyses in pediatric and adult participants yielded a broadly similar pattern.

**Fig. 3.**
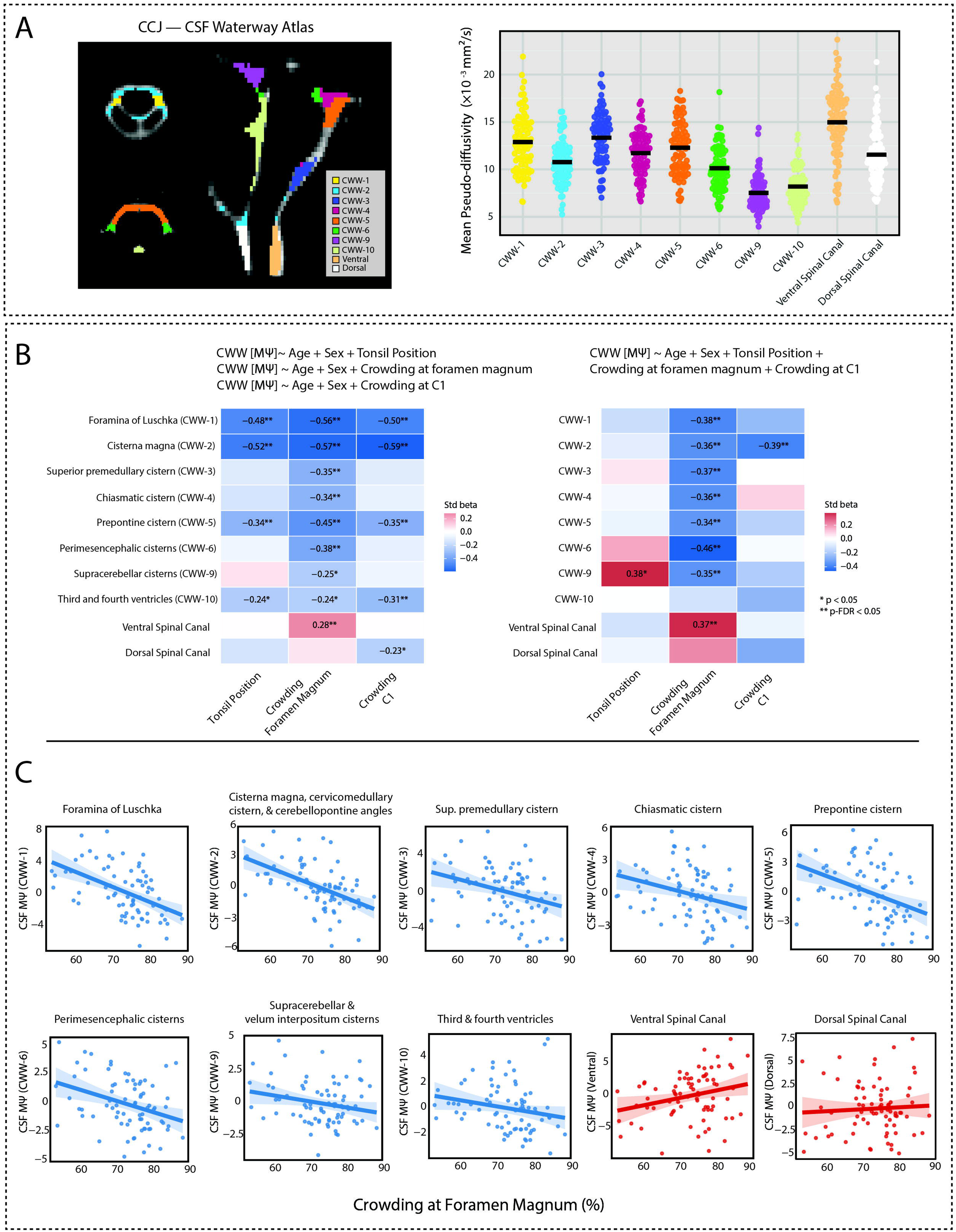
Effects of CCJ structural markers on CSF effective motility across CSF Waterways. **(A)** The CCJ-CSF Waterway atlas: CWW-1, foramen of Luschka; CWW-2, cisterna magna, cervicomedullary cistern, and cerebellopontine angles; CWW-3, superior premedullary cistern; CWW-4, chiasmatic cistern; CWW-5, prepontine cistern; CWW-6, perimesencephalic cisterns; CWW-9, supracerebellar and velum interpositum cisterns; CWW-10, third and fourth ventricles, along with manually delineated ventral and dorsal spinal canals. The distribution of mean pseudo-diffusivity (MΨ) is illustrated across these CWWs and the spinal CSF compartments. **(B)** ROI analyses (n=77) show the effect of CCJ structural markers on regional CSF effective motility across the CWW regions and spinal CSF compartments estimated using linear models adjusted for age and sex. Results are presented for both separate models for each structural marker (tonsillar descent, crowding at both foramen magnum and at C1-level) and a combined model integrating all markers simultaneously. Asterisks (*) indicate significant effects: *p* < 0.05 (*), *p_FDR_* < 0.05 (**). **(C)** Scatter plots illustrate the associations between crowding at the foramen magnum (%) and CSF effective motility across CWW regions, based on the separate linear model adjusted for age and sex. *Abbreviations: CCJ, craniocervical junction; CSF, cerebrospinal fluid; CWW, CSF Waterway; MΨ, mean pseudo-diffusivity; ROI, region-of-interest; FDR, false discovery rate*

However, when tonsil position and CCJ crowding measures were included in the model, the associations between tonsil position and regional CSF MΨ were attenuated and no longer significant (**Fig. 3B**). This attenuation was further supported by mediation analysis, which demonstrated that the effects of tonsil position on reduced CSF MΨ in the basilar cisterns were largely mediated by greater CCJ crowding. For foramen of Luschka (CWW-1) and cisterna magna (CWW-2), the effects of tonsil position were predominantly mediated by crowding at the foramen magnum and C1, with total indirect effects accounting for 85.1% and 73.8% of the total effect, respectively. For CWW-5, the effect of tonsil position was primarily mediated by foramen magnum crowding, with mediation accounting for 83.1% of the total effect **(Supplemental Fig. S2)**. In the combined model, tonsil position was associated with higher CSF MΨ in supracerebellar cistern (CWW-9), in contrast to the effects observed for crowding.

Using multivariate MΨ profiles across crowding-associated CWW regions, we applied MANOVA with Pillai’s trace to identify two foramen magnum crowding thresholds that maximized between-group differences in regional CSF MΨ patterns. The optimal thresholds were 69.5% and 77.5%, defining mild, moderate, and severe crowding groups with robust separation **(Fig. 4A)**. These data-driven groups exhibited graded, region-specific reductions in CWW MΨ with increasing crowding, except for ventral spinal canal CSF, which showed higher MΨ in the intermediate group compared to mild crowding (**Fig. 4B**).

**Fig. 4.**
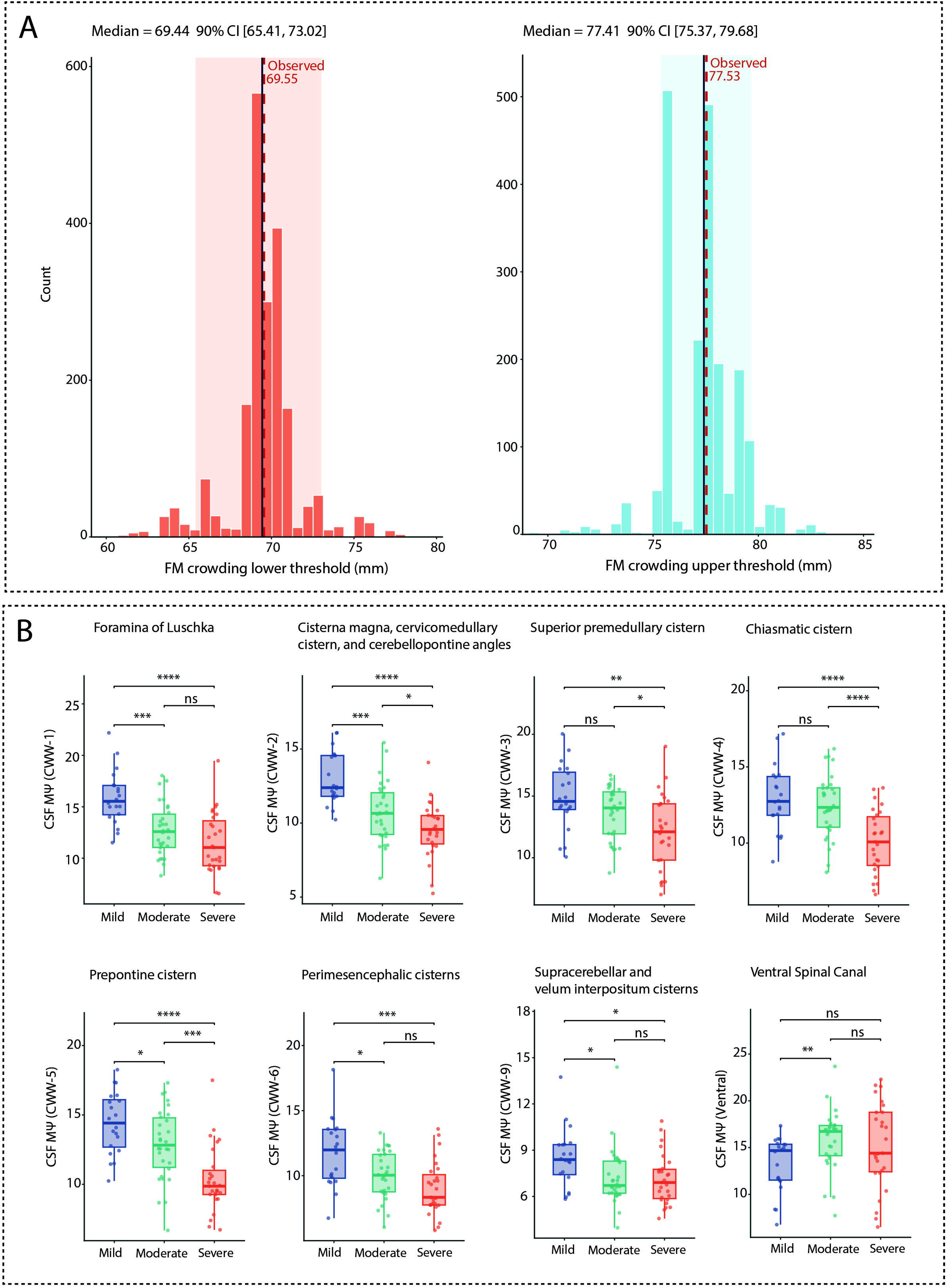
Data-driven stratification of foramen magnum crowding and its association with regional CSF effective motility. **(A)** Distribution of optimal lower and upper thresholds for foramen magnum (FM) crowding derived from bootstrap resampling of multivariate CWW MΨ profiles (n=77). Histograms represent the distribution of threshold values across bootstrap iterations. The dashed vertical lines indicate the observed optimal thresholds (lower: 69.55%; upper: 77.53%), with corresponding median estimates and 95% confidence intervals (lower: median 69.34%, 95% CI [63.57, 75.74]; upper: median 77.45%, 95% CI [70.67, 82.76]), demonstrating stability of the data-driven cut-points. **(B)** ROI analyses showing CSF effective motility (MΨ) across CWW regions stratified by FM crowding severity (mild, moderate, severe) using the data-driven thresholds. Greater crowding is associated with graded reductions in CSF MΨ across intracranial CSF compartments. In contrast, the ventral spinal canal demonstrates a distinct pattern, with relatively increased CSF MΨ in the moderate group. Asterisks (*) indicate significant effects from pairwise t-tests: ns, not significant; *p* < 0.05 (*); *p* < 0.01 (**); *p* < 0.001 (***); *p* < 0.0001 (****). Abbreviations: CCJ, craniocervical junction; CSF, cerebrospinal fluid; CWW, CSF Waterway; FM, foramen magnum; MΨ, mean pseudo-diffusivity; ROI, region-of-interest.

### Syringomyelia and CCJ CSF Effective Motility

Among 81 participants, 21 (25%) had a syrinx (12 cervical). Voxel-wise analyses revealed no significant association between presence of syringomyelia and CSF MΨ. Region-of-interest analyses showed negative associations between CSF MΨ and syringomyelia across multiple CWW regions after adjusting for age and sex; however, these effects were attenuated when further adjusting for crowding and or tonsil position. Only perimesencephalic CWW-6 remained significantly associated with syrinx after adjustment for tonsil position (β = −0.28, p = 0.02; **Supplemental Fig. S3**).

### Comparative Analysis of CSF Flow Dynamics Using MΨand C2–C3 PC-MRI Metrics

We computed correlation coefficients between MΨ values across intracranial and spinal CSF compartments and PC-MRI metrics (n=81) derived from the spinal subarachnoid space at C2–C3 (**Fig. 5A**). Higher stroke volume at C2–C3 was significantly associated with higher MΨ values in the cisterna magna (CWW-2), third and fourth ventricles (CWW-10), supracerebellar cistern (CWW-9), chiasmatic cistern (CWW-4), prepontine cistern (CWW-5), perimesencephalic cisterns (CWW-6), and dorsal spinal regions (p_FDR_ <0.05). Correlations were weaker for peak diastolic and systolic velocities (**Fig. 5B and 5C**). Overall, these findings suggest that stroke volume is more closely related to CWW MΨ, a time-averaged measure of effective CSF motility irrespective of flow direction.

**Fig. 5.**
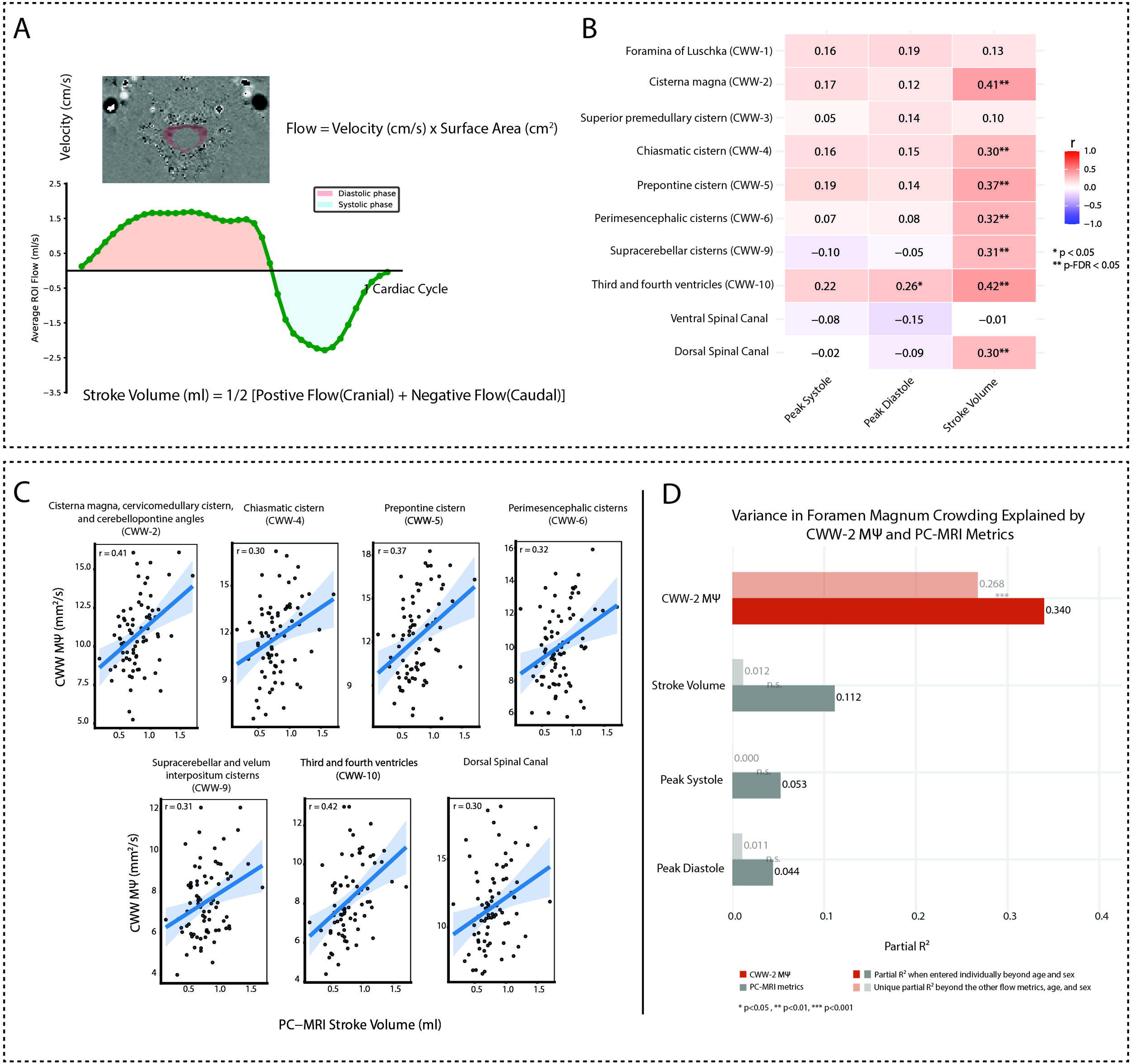
Comparative analysis of CSF dynamics using low-b dMRI (MΨ) and PC-MRI metrics. **(A)** Illustration of PC-MRI–based CSF flow quantification at the C2–C3 level, including velocity–time waveform across the cardiac cycle and derivation of stroke volume. Stroke volume is defined as half the absolute area under the velocity–time curve across the cardiac cycle multiplied by the cross-sectional area of the spinal subarachnoid space at the C2–C3 level. **(B)** Heatmap illustrating the correlations between PC-MRI metrics (peak systole, peak diastole, and stroke volume) and regional CSF MΨ across CWWs (n=81). Stroke volume exhibits the strongest and most widespread associations with CSF effective motility. Asterisks (*) indicate significant effects: *p* < 0.05 (*), *p_FDR_*< 0.05 (**). **(C)** Scatter plots illustrating the linear associations between PC-MRI derived stroke volume and CSF effective motility across CWW regions and spinal CSF compartments. **(D)** CWW-2 MΨ demonstrated the strongest association with crowding, explaining 35.3% of variance after adjustment for age and sex (partial R² = 0.35, p < 0.001) and retaining 28.9% unique variance after accounting for all PC-MRI measures. In contrast, PC-MRI metrics (stroke volume, peak systolic velocity, and peak diastolic velocity) explained substantially less variance individually (partial R² < 0.10) and contributed minimal additional explanatory value once MΨ was included in the model. *Abbreviations: dMRI, diffusion-weighted MRI; CCJ, craniocervical junction; CSF, cerebrospinal fluid; CWW, CSF Waterway; MΨ, mean pseudo-diffusivity; PC-MRI, phase-contrast MRI; ROI, region-of-interest; FDR, false discovery rate*

Next, we compared CWW MΨ and C2–C3 PC-MRI measures in their association with CCJ crowding. Among the CWWs, cisterna magna (CWW-2) MΨ showed the strongest association with foramen magnum crowding, explaining 34% of the variance after adjustment for age and sex (partial R^2^ = 0.34, p < 0.001); accordingly, CWW-2 was selected for comparative analyses. CWW-2 retained 26.8% unique variance after accounting for all C2–C3 PC-MRI flow metrics. In contrast, PC-MRI-derived measures, including stroke volume, peak systolic velocity, and peak diastolic velocity, each explained less than 12% of the variance individually and contributed minimal additional explanatory value (partial R^2^ ≤ 0.01) once CWW-2 was included in the model **(Fig. 5D).**

### Structural correlates of extreme intraventricular MΨoutliers

Six participants exhibited clusters of extreme MΨ values, defined by a voxel-wise z-score threshold exceeding 3.5 and a cluster size greater than 50 voxels; in three of these participants, the high-MΨ clusters were located in the fourth ventricle. Review of structural images revealed a low-lying obex and syringomyelia. Additionally, all three participants exhibited third ventricular enlargement with prominent infundibular and chiasmatic recesses, findings suggestive of an underlying obstructive process affecting intraventricular CSF dynamics **(Fig. 6).** All three features were significantly enriched among fourth ventricular outliers: low-lying obex (3/3 vs. 23/78; p = 0.03), syringomyelia (3/3 vs. 18/78; p = 0.016), and third ventricular abnormalities (3/3 vs 6/78; p < 0.001; all by Fisher’s exact test). Clinically, among these three participants, one had symptoms and signs of bulbar dysfunction including hoarseness, dysphagia, choking, and vocal cord dysfunction; two had sleep apnea, and two had urinary incontinence. Gag-reflex data were available for two participants, both of whom had a diminished gag reflex. None of these clinical features was more frequent among fourth-ventricular outliers than among the remaining participants. Two additional participants with outlier clusters in the supracerebellar and quadrigeminal cisterns demonstrated borderline tonsillar ectopia with minimal foramen magnum crowding and no other structural abnormalities.

**Fig. 6.**
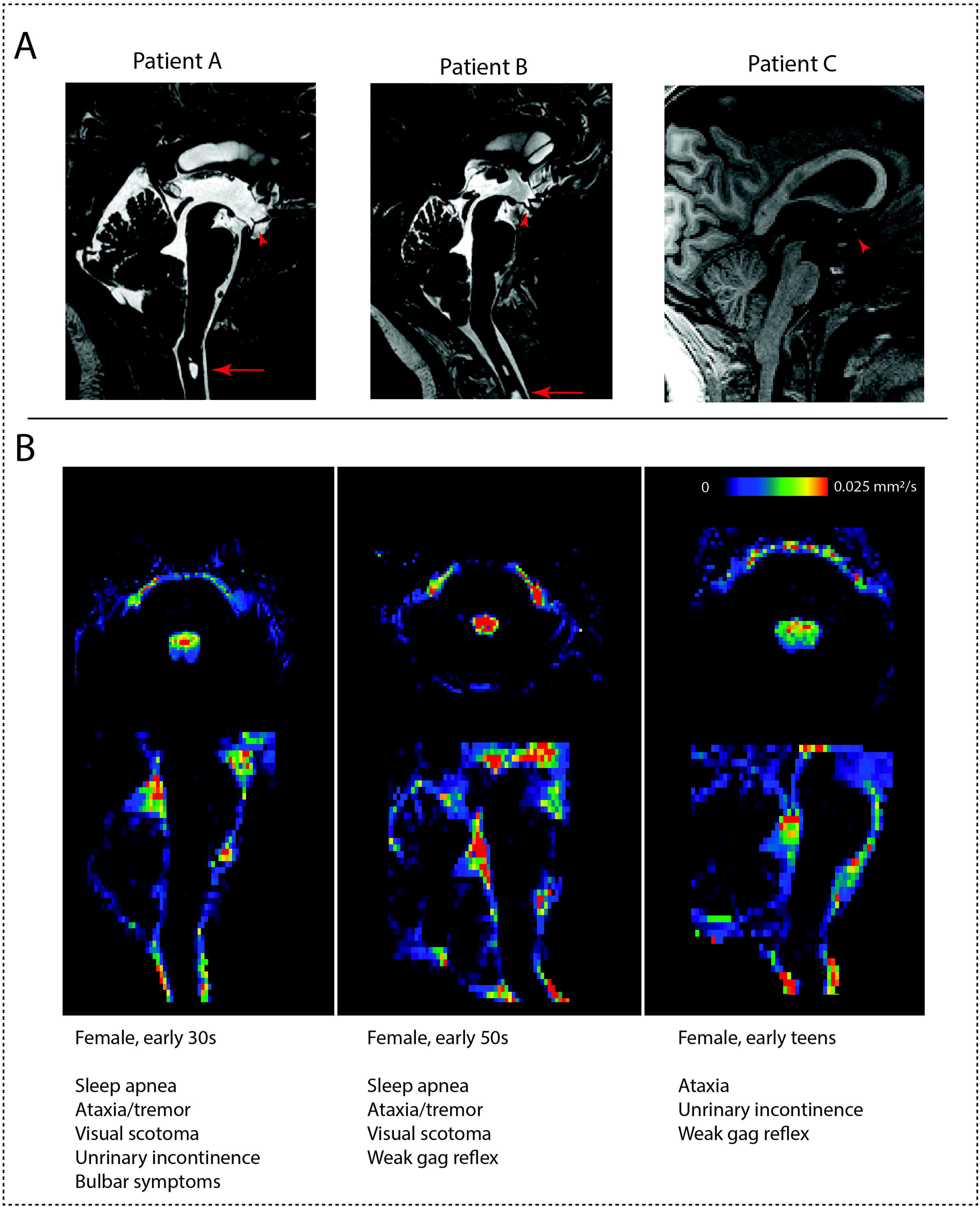
Structural abnormalities associated with extreme intraventricular CSF effective motility. **(A)** Mid-sagittal structural MRI images from three participants exhibiting extreme CSF MΨ values within the fourth ventricle. Red arrowheads indicate prominent infundibular and chiasmatic recesses, consistent with third ventricular enlargement. Arrows denote syringomyelia in patients A and B; in patient C, the syrinx is outside the field of view. **(B)** Corresponding low b-value dMRI-derived MΨ maps in axial and sagittal views demonstrate focal clusters of increased CSF effective motility within the fourth ventricle. One participant exhibited bulbar symptoms including hoarseness, dysphagia, choking, and vocal cord dysfunction. *Abbreviations: CSF, cerebrospinal fluid; MΨ, mean pseudo-diffusivity; dMRI, diffusion-weighted MRI*

### Baseline CSF Motility and Its Associations with Patient-Reported Outcomes and Subsequent Decompression Surgery

Lower baseline CSF MΨ was associated with worse patient-reported outcomes on PROMIS after adjusting for age and sex. Among the 50 adult subjects with available baseline PROMIS data (mean age: 34.5 ± 11.4), greater pain interference was associated with lower CSF MΨ in the superior premedullary cistern (CWW-3, β *=* -0.3, *p* = 0.03), perimesencephalic cisterns (CWW-6, β *=* -0.4, *p* = 0.005) and supracerebellar interpositum cisterns (CWW-9, β *=* -0.32, *p* =0.04). Lower CSF MΨ was associated with reduced cognitive function within the supracerebellar cistern (CWW-9, β *=* 0.43, *p* = 0.004) and perimesencephalic cisterns (CWW-6, β *=* 0.3, *p* = 0.04; **Fig. 7A**). CHIP analyses were limited to 25 pediatric patients with available baseline scores (mean age: 12.8 ± 2.59 years). No significant association was observed between CWW MΨ and CHIP composite score or any of its subdomains.

**Fig. 7.**
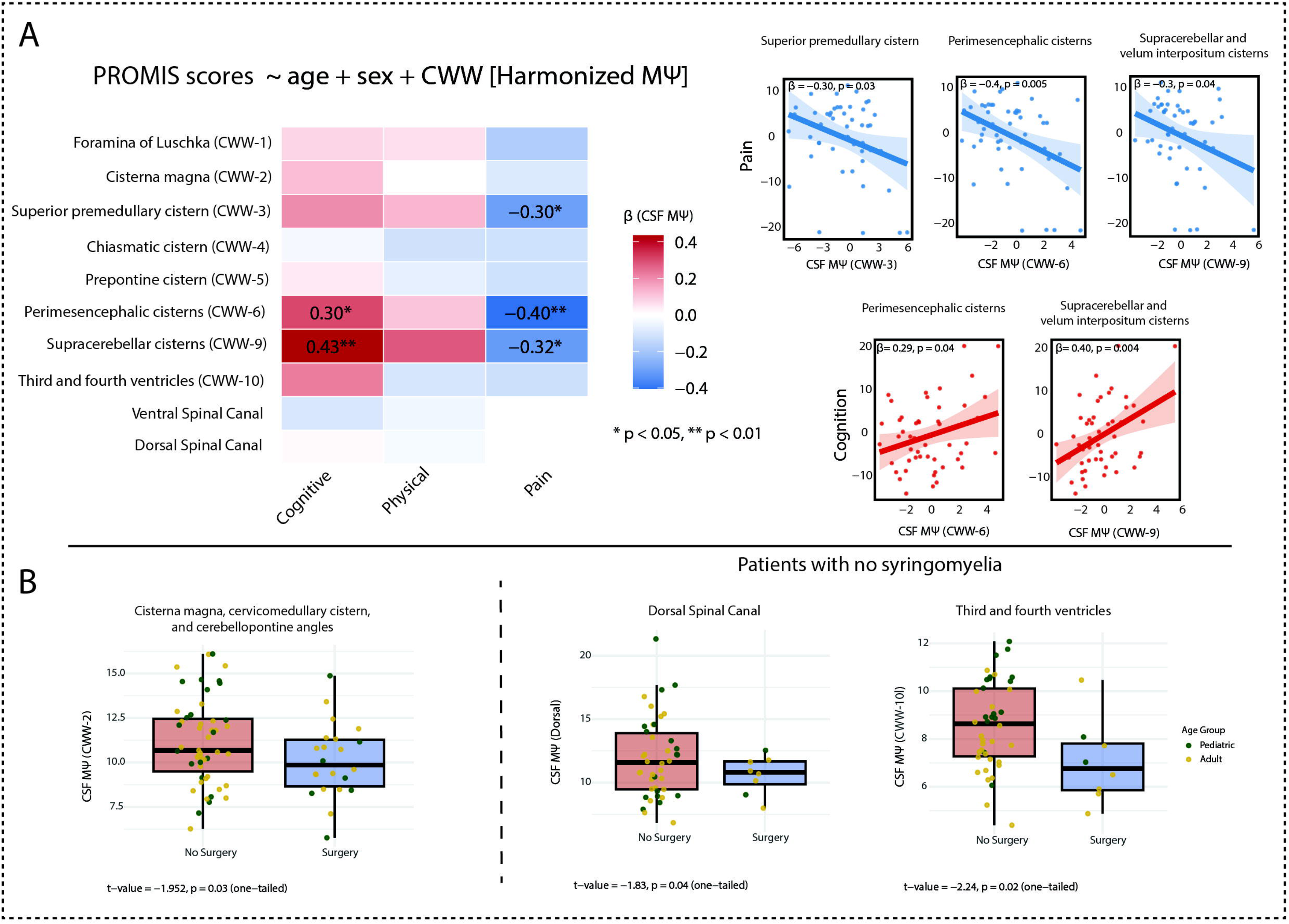
Clinical associations and surgical status related to CSF effective motility. **(A)** ROI analyses (n=50) show the association between regional CSF effective motility and PROMIS T-scores (pain interference, physical function and cognitive function) in adults adjusted for age and sex. Lower effective motility within the superior premedullary (CWW-3), perimesencephalic (CWW-6), and supracerebellar (CWW-9) cisterns is associated with greater pain interference. Additionally, lower motility in the perimesencephalic and supracerebellar cisterns is linked to reduced cognitive function. Asterisks (*) indicate significant effects: *p* < 0.05 (*), *p* < 0.01 (**). **(B)** Box plots illustrate the baseline CSF MΨ levels of subjects who subsequently underwent decompression surgery compared to those who did not. Patients who received surgery demonstrated significantly lower preoperative motility in the cisterna magna (CWW-2). Among subjects without syringomyelia, lower baseline motility in the third and fourth ventricles (CWW-10) and the dorsal spinal canal was also observed in those who underwent surgery. *Abbreviations: CSF, cerebrospinal fluid; CWW, CSF Waterway; MΨ, mean pseudo-diffusivity; PROMIS, Patient-Reported Outcomes Measurement Information System*.

Among 73 subjects with available surgical status, exploratory analysis revealed that patients who underwent decompression surgery (n=22) demonstrated lower pre-operative CSF MΨ within cisterna magna (CWW-2) compared to those who did not undergo surgery (10.1 ± 2 vs. 11.2 ± 2.4; t = −1.95, one-tailed *p* = 0.02). Given that surgical decision-making is often influenced by the presence of a syrinx, we next repeated the analysis after excluding subjects with syringomyelia. Among the remaining 50 subjects, of whom 8 underwent decompression surgery, lower baseline CSF MΨ in the surgical group was observed in the third and fourth ventricles (CWW-10: 7 ± 1.75 vs. 8.5 ± 1.8; t = −2.24, p = 0.02) and dorsal spinal regions (10.6 ± 1.5 vs. 11.9 ± 3.1; t = −1.83, p = 0.04; all one-tailed; **Fig. 7B**).

## Discussion

In this study, we developed a high-resolution low-b dMRI framework to map CSF effective motility across the CCJ, upper cervical spine, and posterior fossa in patients with CM-I. By combining reduced field-of-view imaging, voxel-wise CCJ-CΨSS analysis, and region-of-interest assessment with the CWW atlas, we showed that CCJ crowding, particularly at the foramen magnum, is more closely linked to widespread reductions in posterior fossa CSF motility than tonsil position alone. The apparent effects of tonsil position on CSF motility were largely attenuated after accounting for CCJ crowding. Mediation analyses further demonstrated that this relationship was largely explained by CCJ crowding.

Specifically, greater crowding at the foramen magnum was associated with widespread reductions in CSF MΨ across ventral basilar cisterns and fourth ventricular outflow pathways, whereas crowding at C1 and tonsil position showed more localized effects near the interpeduncular cistern and fourth ventricular outflow pathways. Obstruction of fourth ventricular outflow is a recognized feature of CM-I and has been proposed as a marker of disease severity^1,6,11^. While prior studies have primarily focused on the foramen of Magendie using mid-sagittal phase-contrast MRI—where velocity encoding is typically applied in the craniocaudal direction—our analysis revealed that CM-I structural severity was associated with reduced MΨ not only at the foramen of Magendie but also at the foramina of Luschka. The oblique orientation of Luschka outflow may limit its examination with conventional PC-MRI, whereas CΨSS enables detection of CSF motion irrespective of flow direction along these lateral pathways. While the prepontine cistern serves as the primary conduit for CSF circulation between the intracranial and cervical compartments^45,46^, few PC-MRI studies have examined CSF dynamics within the basilar cisterns. Notably, one report described increased flow velocities in the cerebellomedullary cistern, adjacent to the outflow region of the foramina of Luschka, following decompressive surgery^47^. This is consistent with our finding that more severe CM-I features are associated with reduced CSF motion in the cisterna magna and adjacent fourth ventricular outflow pathways.

Greater crowding at the foramen magnum was also associated with elevated CSF MΨ within the ventral upper cervical spinal canal. This finding is consistent with prior PC-MRI studies demonstrating heightened flow velocities, localized flow jets, and complex CSF flow patterns within the ventral spinal canal in CM-I^16,17,48,49^. Computational fluid dynamics studies suggest that crowding at the CCJ increases impedance to CSF motion and amplifies pressure gradients across the foramen magnum. During systole, CSF is funneled through narrow ventral pathways across the CCJ, generating flow jets and complex flow patterns at the foramen magnum and within the ventral upper cervical spinal canal^4,45,50–52^. A possible explanation for the divergent effects of crowding at the foramen magnum on intracranial versus spinal CSF dynamics is a systole-predominant bottleneck effect at the CCJ. During systole, active expansion of intracranial vasculature generates a strong, transient cranio-spinal pressure gradient, producing a sharp caudally directed CSF pulse from the prepontine cistern into the cervical subarachnoid space^53^.

In CM-I, crowding at the foramen magnum may impede this systolic displacement proximally, reducing CSF effective motility in the prepontine cistern. At the same time, the same pressure gradient may force CSF through narrow residual ventral pathways across the crowded foramen magnum, producing high-velocity jets and elevated MΨ in the downstream ventral upper cervical spinal canal. In contrast, the slower, longer-duration diastolic CSF flow may lack the transient pressure impulse needed to generate jet-like acceleration across the crowded foramen magnum. Instead, increased impedance at the CCJ may simply attenuate diastolic CSF velocity, resulting in dampened CSF motion within the basilar cisterns.

A common site of decreased MΨ associated with both greater tonsillar descent and CCJ crowding was the interpeduncular cistern, where the Liliequist membrane is located^54,55^. This likely reflects the role of arachnoid membranes in shaping and dispersing CSF flow. In more severe CM-I, the flow shaping and dispersive effects of these arachnoid membranes may be diminished by increased impedance and reduced diastolic flow. These findings align with our prior work demonstrating that fine arachnoid structures can shape intracranial CSF flow patterns^21^. Furthermore, discrepancies between CFD simulations and 4D phase-contrast MRI have been attributed to such subtle anatomical features, underscoring their physiological significance^45,56^. Taken together, ultra-high resolution structural imaging could help delineate these fine arachnoid membranes and further elucidate the structural basis of CSF flow abnormalities in CM-I.

While cerebellar tonsillar descent remains the defining imaging feature of CM-I^1^, our findings underscore that it captures only one aspect of the pathophysiology. It has been postulated that the rigid osseous ring of the foramen magnum contributes to impaired accommodation of cerebellar herniation and CSF pulsatility, leading to dynamic subarachnoid space compression, reduced intracranial compliance, and syringomyelia formation^57,58^. Mediation analysis showed that the association between tonsil position and intracranial CSF flow was largely accounted for by CCJ crowding, suggesting that impaired CSF motion is more closely related to structural crowding than to tonsil position itself. Consistent with these findings, prior work has shown that foramen magnum crowding is a stronger predictor of symptomatology and surgical intervention than tonsillar descent^59^. Although foramen magnum and C1 crowding both reflect structural narrowing at the CCJ, their only moderate correlation and differing spatial associations with CSF motility suggest that they capture complementary aspects of CCJ obstruction. Similarly, a prior CFD study showed that elevated CSF flow impedance was only partially explained by tonsillar descent, with additional contributions from CCJ crowding, narrowed subarachnoid spaces, and other structural factors not captured by tonsillar ectopia alone^4^. These findings collectively point to a multifactorial model of CSF flow impairment in CM-I, wherein distinct morphometric features interact to modulate neurofluid dynamics^3^.

We observed that the presence of a syrinx was associated with reduced CSF motility across multiple regions; however, these associations were largely attenuated after adjustment for structural crowding or tonsillar descent. This suggests that the association between syringomyelia and impaired CSF motility primarily reflected their shared relationship with CM-I severity at the CCJ, rather than an independent effect of syringomyelia. This is consistent with prior work demonstrating association between syrinx and structural crowding at foramen magnum^60^. However, given the small subsample with syrinx in this study (n = 21), larger studies are needed to more comprehensively characterize how syrinx location (cervical versus thoracic) and morphological features, including degree of expansion (markedly expansile versus small or slit-like cavities), influence CSF dynamics.

Among the PC MRI-derived CSF flow metrics, stroke volume exhibited stronger correlations with low b-value dMRI-derived MΨ than the peak velocities. This likely reflects their shared characterization as time-averaged, non-directional measures, and also the fact that higher CSF stroke volume at C2–C3 is expected to influence inflow–outflow dynamics and overall CSF motion within the CCJ. In contrast, peak velocity metrics, while widely reported, have shown inconsistent findings across prior studies, as they vary by phase and region and are more susceptible to noise, aliasing, and confounding from adjacent vascular flow^16,61^. Moreover, the two approaches have distinct sensitivities: conventional 2D PC-MRI is primarily sensitive to bulk, craniocaudal CSF flow driven by the cardiac cycle, whereas low-b dMRI captures more complex, multidirectional motion (including mixing and vortical patterns)^18,20^ that are particularly prominent in CM-I^48^. These findings underscore the complementary nature of low b-value dMRI and PC MRI in characterizing CSF dynamics, with each providing unique sensitivity to distinct aspects of CSF motion. CWW MΨ emerged as the dominant correlate of CCJ crowding, capturing substantially greater and largely independent variance compared with C2–C3 PC-MRI metrics, which contributed minimal additional explanatory value. These findings highlight a key limitation of conventional 2D PC-MRI, namely its reliance on predefined slice positioning. Measurements at a fixed level such as C2–C3 may not capture the spatially localized and heterogeneous CSF flow alterations at the CCJ in CM-I, contributing to their limited explanatory power relative to CWW MΨ. 4D flow PC-MRI, with three-dimensional spatial coverage, may therefore provide a more comprehensive assessment of CSF dynamics across the CCJ.

Our work has immediate clinical implications. First, the data-driven thresholds for foramen magnum crowding could be readily applied for objective radiological grading of CM-I, providing a physiologically informed framework that complements conventional structural assessment. In addition, extreme fourth ventricular MΨ elevations were observed in three participants and were accompanied by syringomyelia, a low-lying obex, and third ventricular enlargement. Together, these findings suggest a CM-I endotype characterized by hyperdynamic fourth ventricular CSF motility, potentially reflecting underlying fourth ventricle outflow obstruction. These cases illustrate that readily identifiable abnormalities on MΨ maps, even by visual inspection, may reflect underlying pathophysiologic processes and highlight the potential for qualitative assessment to inform routine radiologic practice. Notably, generating MΨ maps requires low-b dMRI with only a limited number of diffusion-encoding directions (as few as 6–20), which can be acquired in 1 to 3 minutes using standard MRI platforms and clinically feasible protocols^21^. In contrast to PC MRI, low-b dMRI does not require VENC calibration, subject-specific parameter adjustment, or selection of flow-sensitization directions, thereby simplifying clinical implementation and reducing operator dependence.

Moreover, our work establishes the clinical significance of CSF effective motility by linking it to patient-reported outcomes in adults. Specifically, lower CSF MΨ was associated with worse pain interference and reduced cognitive function, suggesting that impaired CSF mixing and circulation may contribute to the chronic symptoms often reported by CM-I patients^62^. However, such associations were not found in the pediatric cohort using CHIP scores, which may reflect the inherent difficulty in capturing quality-of-life metrics in children or the naturally heterogenous clinical spectrum of CM-I^63,64^. Building on this association between CSF flow and symptoms, our exploratory analyses suggest that baseline CSF motility may also serve as a functional biomarker for surgical candidacy. Patients who eventually required decompression surgery demonstrated lower preoperative CSF motility in the cisterna magna, third and fourth ventricles and dorsal spinal regions. This indicates that low-b dMRI can identify physiological flow impairments that warrant intervention. In line with these findings, recent work has shown that presurgical measures of CSF flow and brain motion are more descriptive of postsurgical dynamic improvements than conventional measures of tonsillar descent^7^

Several limitations should be acknowledged. While the sample size was sufficient to evaluate the effects of CM-I structural severity on CSF flow, larger studies are needed to better characterize the heterogeneity of CSF flow abnormalities in CM-I, including the impact of major CM-I complications such as syringomyelia and hydrocephalus. In this study, low-b dMRI acquisitions were not physiologically gated; as such, MΨ values reflect an aggregate measure of effective CSF motility rather than time-resolved dynamics. Recent work suggests that gated low-b dMRI can capture CSF flow variations across the cardiac cycle, offering additional physiological insights^22,65^. Furthermore, MΨ was computed over a volumetric region encompassing the spinal canal from the CCJ to the C1–C2 levels, whereas PC-MRI metrics were derived from a single axial slice at the C2–C3 level, limiting the direct comparability between modalities. A head-to-head evaluation will require volumetric 4D flow acquisitions to match the spatial extent and multi-directional sensitivity of MΨ mapping, enabling more rigorous cross-validation of CSF motion features across techniques.

This study demonstrates that low-b dMRI coupled with CΨSS workflow, provides a comprehensive and spatially unbiased map of CSF flow impairment in CM-I. Our results underscore that CCJ crowding is the primary structural determinant of impaired CSF effective motility, superseding tonsillar descent. An integrated structure–function assessment of CCJ anatomy and CSF dynamics represents a key direction for advancing patient-specific evaluation and management in CM-I.

## Data Availability

The data that support the findings of this study were generated as part of the ongoing Redefining Chiari Type I Malformation and its Impact on Brain Development prospective cohort study at Washington University School of Medicine in St. Louis. The data are not publicly available because they contain human participant information and are subject to institutional and privacy restrictions. De-identified data may be made available upon reasonable request to the corresponding author and with appropriate institutional approvals.

## Data and Code Availability

The CCJ-CΨSS framework will be made available as open-source software at https://github.com/brafti, with accompanying template files deposited in an open research repository.

---

**Supplemental Fig. S1. Overview of the Craniocervical Junction (CCJ) CSF Pseudo-diffusion Spatial Statistics (CΨSS) workflow. (1)** Low b-value dMRI (multi-b-value subset: b = 0–350 s/mm^2^, 29 diffusion-weighted volumes, 7 b0 volumes; b ≥ 650 s/mm^2^, 15 diffusion-weighted volumes) were utilized to generate low b-value MΨ maps, measuring CSF effective motility, which captures a spectrum of flow regimes including laminar, vortical, and mixing patterns. **(2)** Study-specific CCJ template was constructed using MΨ and pseudo-T1 (generated via Atropos two-tissue segmentation) as multivariate inputs for *antsMultivariateTemplateConstruction2.sh* (ANTs). Subsequently, individual MΨ and CSF partial volume maps were registered to this template space. A final CSF mask was generated by retaining voxels with a CSF fraction > 0.8 in at least 75% of subjects. Voxels with a lower CSF fraction were filled using surrounding voxel averages. Spatial smoothing (3 mm FWHM) was applied within the CSF mask using 3dBlurInMask (AFNI).

*Abbreviations: CSF, cerebrospinal fluid; CCJ, craniocervical junction; FWHM, full width at half maximum; dMRI, diffusion-weighted MRI; MΨ, mean pseudo-diffusivity*.

**Supplemental Fig. S2. Craniocervical junction crowding mediates the association between tonsil position and regional CSF effective motility.** Parallel mediation analyses evaluating whether FM and C1 crowding mediate the association between tonsil position and MΨ in **(A)** the foramina of Luschka outflow (CWW-1), **(B)** the cisterna magna (CWW-2), and **(C)** the prepontine cistern (CWW-5). Direct, indirect, and total effects are presented as standardized regression coefficients (β), with corresponding 95% confidence intervals (CI) and *p* values. All models were adjusted for age and sex.

*Abbreviations: CSF, cerebrospinal fluid; FM, foramen magnum; MΨ, mean pseudo-diffusivity; CWW, CSF Waterway*.

**Supplemental Fig. S3. Association between syringomyelia and regional CSF effective motility.** Heatmap illustrating the associations between syringomyelia and regional CSF effective motility (MΨ) across CWW regions under three regression models. The base model was adjusted for age and sex, Model 1 additionally adjusted for tonsil position, and Model 2 additionally adjusted for foramen magnum (FM) crowding. β coefficients are shown for each model, with asterisks (*) indicating significant effects (*p* < 0.05).

*Abbreviations: CSF, cerebrospinal fluid; CWW, CSF Waterway; FM, foramen magnum; MΨ, mean pseudo-diffusivity*.

