## Supplementary figures and images for "Moving Past Tonsil Position: Craniocervical Junction Crowding Shapes Cerebrospinal Fluid Effective Motility in Chiari I Malformation"

### Supplementary Figure S1

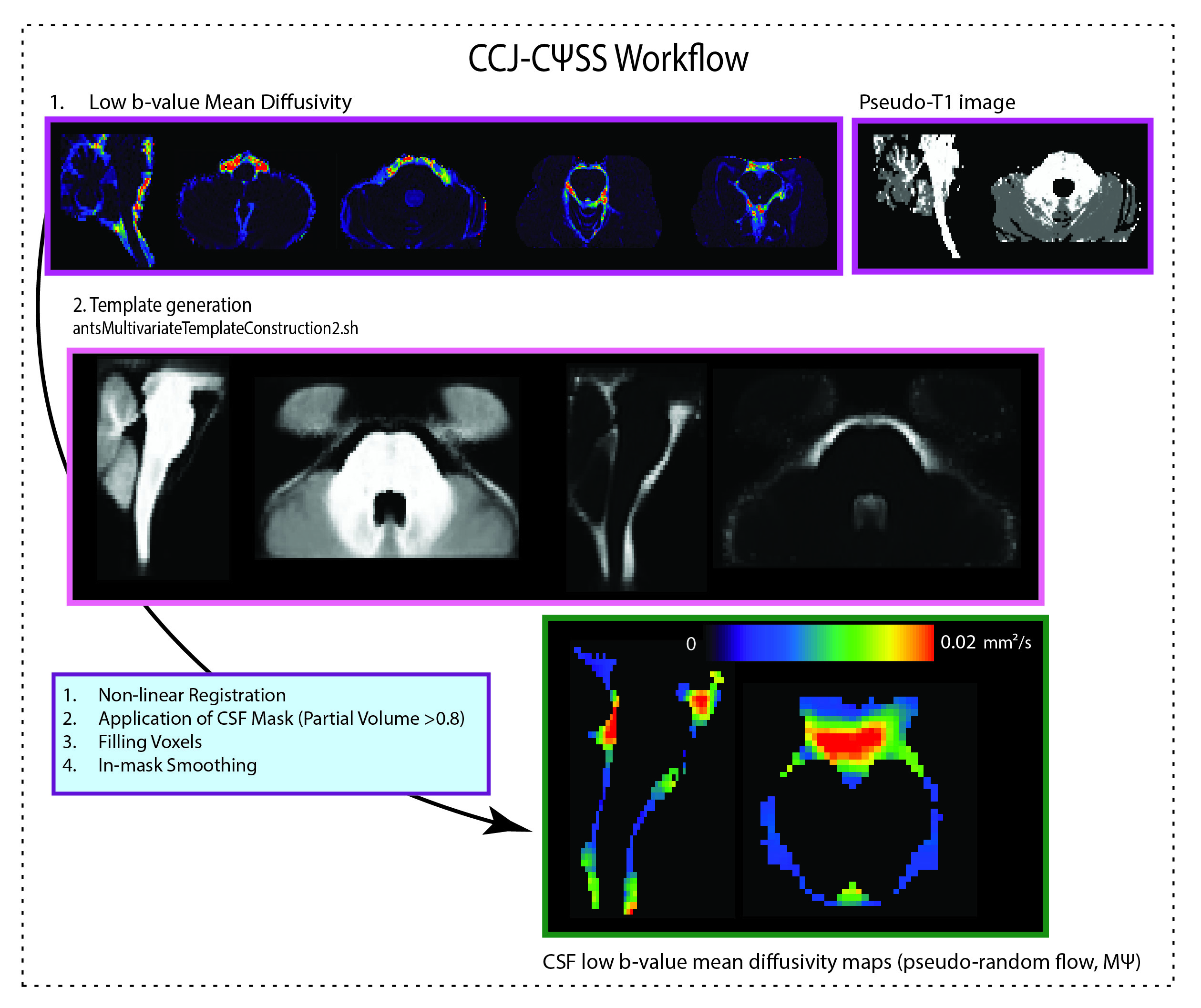

### Supplementary Figure S2

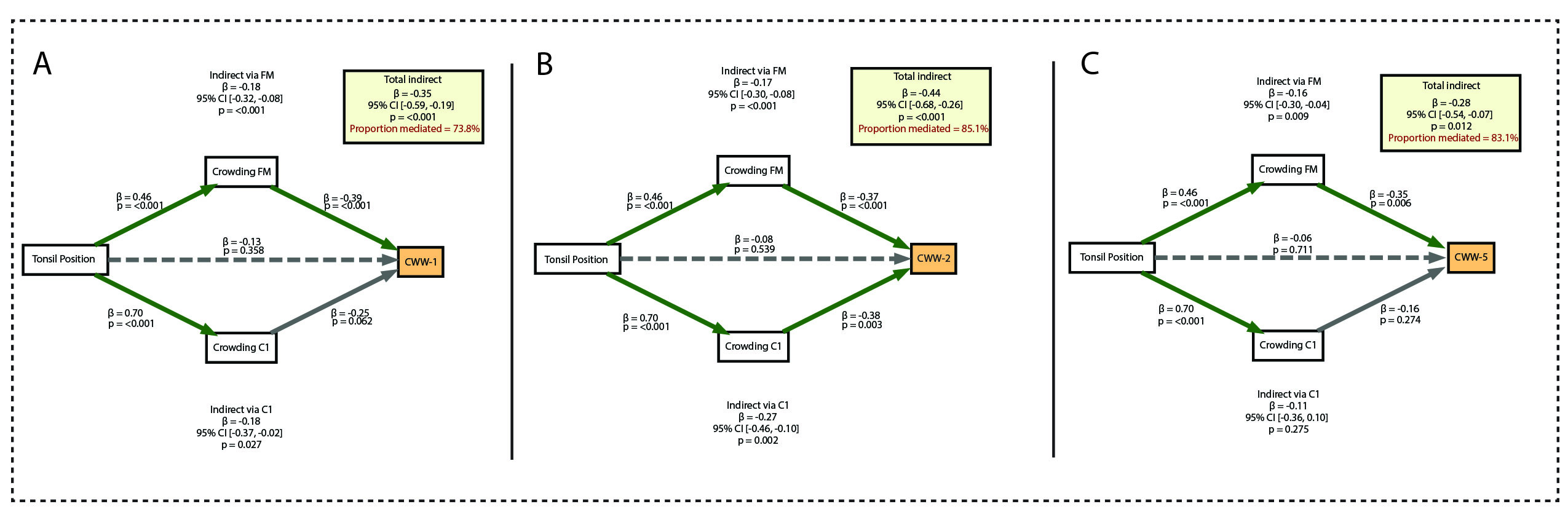

### Supplementary Figure S3

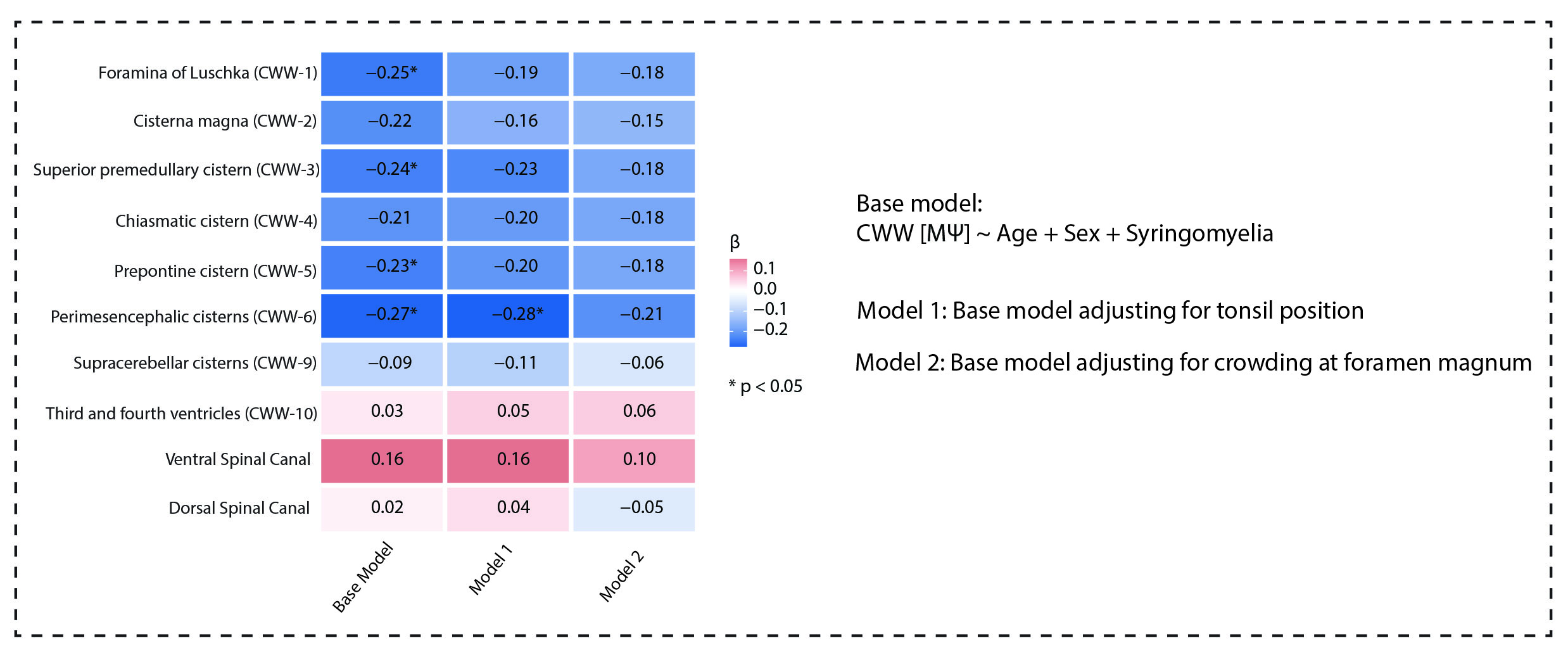
